# Sex differences in COVID-19 infection and mortality in Hong Kong

**DOI:** 10.64898/2026.03.07.26347844

**Authors:** Alexandra H.T. Law, Jessica Y. Wong, Yun Lin, Benjamin J. Cowling, Peng Wu

## Abstract

**Background:** Variation in COVID-19 mortality rates by sex could have several explanations. We aimed to determine sex differences in infection and mortality patterns across different COVID-19 epidemics in Hong Kong, and to evaluate potential hypotheses.

**Methods:** We estimated age- and sex-specific incidence rates of cases, hospitalizations, and deaths per 100,000 population. Case-hospitalization, case-fatality risks (CFRs), and hospital-fatality risks of the COVID-19 pandemic were also estimated. Adjusted and unadjusted risks were estimated and compared to explore the relationships between mortality and health-related variables. We also explored the sex ratio of COVID-19 mortality rates of respiratory diseases from 2000 to 2019.

**Results:** Hong Kong recorded 2876110 COVID-19 cases and 12737 deaths between January 2020 and January 2023, with 1317368 cases (45.8%) and 7523 (59.1%) fatal cases occurring in males. The incidence rate of cases was similar by sex across waves. The CFRs and hospital-fatality risks were higher in men across all waves. Males had a significantly higher mortality risk after adjusting for sex, COVID-19 vaccination status, and pre-existing chronic diseases. The ratio of COVID-19 mortality rates in men versus women from 2020 to 2023 was similar to the mortality ratio for other respiratory diseases in the pre-pandemic period.

**Conclusions:** While infection rates were similar for males and females, males experienced higher mortality risks even after adjusting for differences in other known risk factors. COVID-19 shares a similar sex ratio of mortality with respiratory diseases excluding COVID-19.

## INTRODUCTION

Several studies have indicated that men experience greater COVID-19 mortality rates than women (1,2), as reflected in aggregate data from various countries. Higher mortality risks were observed in males in the United States (3,4), the Netherlands (5), Peru (6), and England & Wales (7), attributed to biological factors, i.e. females develop higher innate and adaptive immunological responses than males because the X chromosome has more immune-related genes, and increased exposure of males to COVID-19 in workplace settings (8). However, Sha et al. (9) found no significant sex difference in in-hospital mortality for the population aged 55 or below in mainland China in 2020. Aleksanyan and Weinman (10) reported that women’s lower access to healthcare resources in developing countries, due to social norms and financial constraints, might have resulted in greater disadvantages for females regarding COVID-19 mortality.

In Hong Kong, with a developed and widely-accessible healthcare system, the initial spread of SARS-CoV-2 was effectively constrained for two years, before the spread of Omicron subvariants resulted in substantial mortality in 2022, predominantly among older age groups (11). Further analysis is required to explore the relationship between COVID-19 mortality and variables such as sex and underlying chronic conditions. This analysis aims to investigate sex differences in COVID-19 incidence and mortality across various epidemic waves and among different age groups. We also examine factors potentially explaining differential mortality rates by sex.

## METHODS

### COVID-19 cases

We obtained individual patient data on COVID-19 cases confirmed from 23 January 2020 to 29 January 2023, including age, sex, history of chronic diseases, date of report, disease outcomes (discharge or death), date of discharge or death, and SARS-CoV-2 vaccination status. All COVID-19 cases were classified as mild, moderate, severe, critical, or fatal according to ultimate clinical outcomes (12). The criteria for case classification are provided in Table S1 of the appendix. Table S2 shows the sex-specific proportions of different conditions defined by the proposed criteria in each wave of the pandemic.

### COVID-19 waves in Hong Kong

We classified the COVID-19 pandemic in Hong Kong into eight waves by the report date of cases, as shown in Table S3 in the appendix. We combined the analysis for the first four waves, which were small in size and occurred with ancestral strains of the virus (not variants) before the availability of vaccines or antivirals (11).

### Sex differences in COVID-19 incidence and fatality

To characterize the sex differences in COVID-19 severity and death over 8 epidemic waves in Hong Kong, we estimated sex-specific numbers of cases, hospitalizations, and deaths per 100,000 population, as well as case-hospitalization rate, case-fatality rate (CFR), and hospital mortality rate for adults aged 45 or above. Wilson score interval was employed to account for statistical uncertainty in estimates, as it performs better than the normal approximation for some age groups that had a small number of deaths in specific waves. Within each wave, the CFR for each age and sex was estimated via the number of deaths divided by the number of confirmed COVID-19 cases in that group (Appendix Table S4). Hong Kong required all confirmed cases to be isolated in hospitals from wave 1 to 4. This policy changed during the fifth wave as case numbers increased substantially and beyond the capacity of hospital isolation facilities. Since hospitalization was a marker of isolation rather than necessarily a marker of severity, we defined hospitalized cases as those cases classified as having either severe or critical or fatal status, to provide a more stable comparison across all waves (13), i.e. not considering those patients isolated with mild disease as being “hospitalized” for the purpose of these analyses. A Wilson score interval was used to construct uncertainty intervals for the rates. The wave-specific number of cases, hospitalizations, and deaths per 100,000 population was calculated by the number of cases, hospitalizations, or deaths divided by the relevant mid-year population size from the Hong Kong Statistics & Census Department (14), which were provided in Table S5.

Logistic regression was conducted to examine the relationship between sex, comorbidities, vaccination status, and mortality while controlling for potential confounders, among confirmed and hospitalized cases. The adjusted and unadjusted odds ratios (ORs) were estimated and compared across adults aged 45 years or above. All analyses were conducted in R version 4.3.3 (R Foundation for Statistical Computing, Vienna, Austria). Our study received ethical approval from the Institutional Review Board of the University of Hong Kong.

### Role of the funding source

The funding bodies had no role in the design of the study, the collection, analysis, and interpretation of data, or writing of the manuscript.

## RESULTS

Figure 1 illustrates daily sex-specific COVID-19 cases per 1,000 population and deaths per 100,000 population from January 23, 2020, to January 29, 2023. Hong Kong effectively contained the spread of COVID-19 during the first two years of the pandemic (waves 1-4). However, a substantial epidemic of the Omicron BA.2 subvariant occurred in early 2022. In wave 5, both daily mortality rates and case numbers peaked for both sexes, with males experiencing a mortality rate of 152.1 deaths per 100,000 population, compared to 86.0 for females (Table 1). Infection rates and mortality rates were lower in waves 6 and 7. However, in wave 8, rates slightly increased: among males, there were 127.8 cases per 1,000 and 46.1 deaths per 100,000, while among females, there were 134 cases per 1,000 and 29.5 deaths per 100,000 (see Table 1).

**Figure 1:**
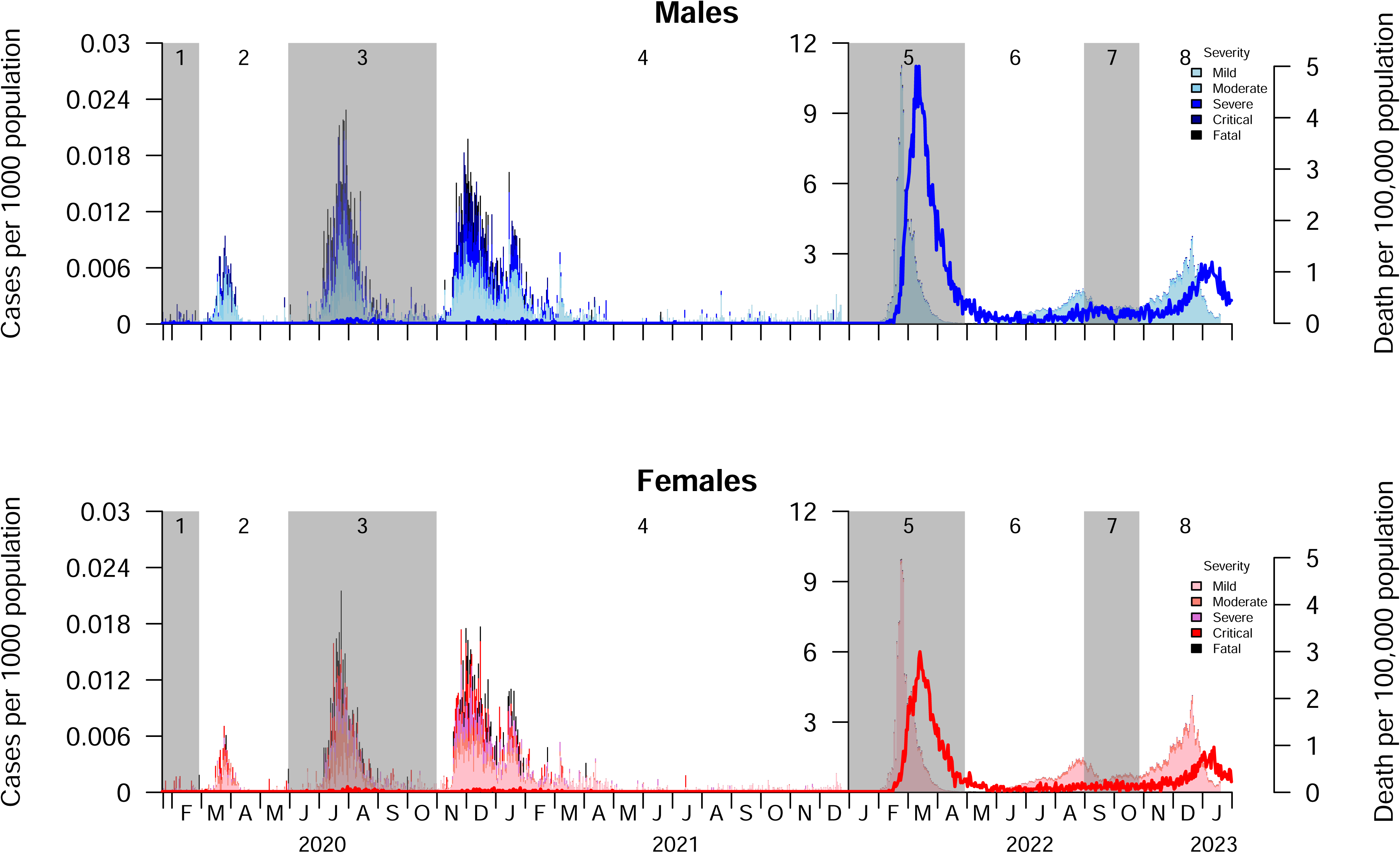
The daily COVID-19 cases per 1000 population and COVID-19 deaths per 100,000 population, by sex.

**Table 1:** The incidence of COVID-19 cases per 1000 population, and COVID-19 mortality rate per 100,000 population, by sex and epidemic waves.

| Wave | Males |  | Females |  |
| --- | --- | --- | --- | --- |
|  | Cases per 1000 population | Deaths per 100,000 population | Cases per 1000 population | Deaths per 100,000 population |
| 1-4 | 1.80 | 3.75 | 1.62 | 2.16 |
| 5 | 166.35 | 152.09 | 156.89 | 85.98 |
| 6 | 45.57 | 10.94 | 43.60 | 6.94 |
| 7 | 48.72 | 10.46 | 49.30 | 5.41 |
| 8 | 127.77 | 46.08 | 134.03 | 29.47 |
| **Overall** | **395.26** | **224.27** | **390.51** | **133.13** |

Figure 2 shows the sex-, age-, and wave-specific number of cases, hospitalizations, and deaths per 100,000 population. Three indicators reached the peak in wave 5 for all age groups. Both sexes experienced similar infection rates across all waves, but males experienced higher hospitalization and deaths than women in all waves: severity increased in wave 8, but is still far from peak in wave 5.

**Figure 2:**
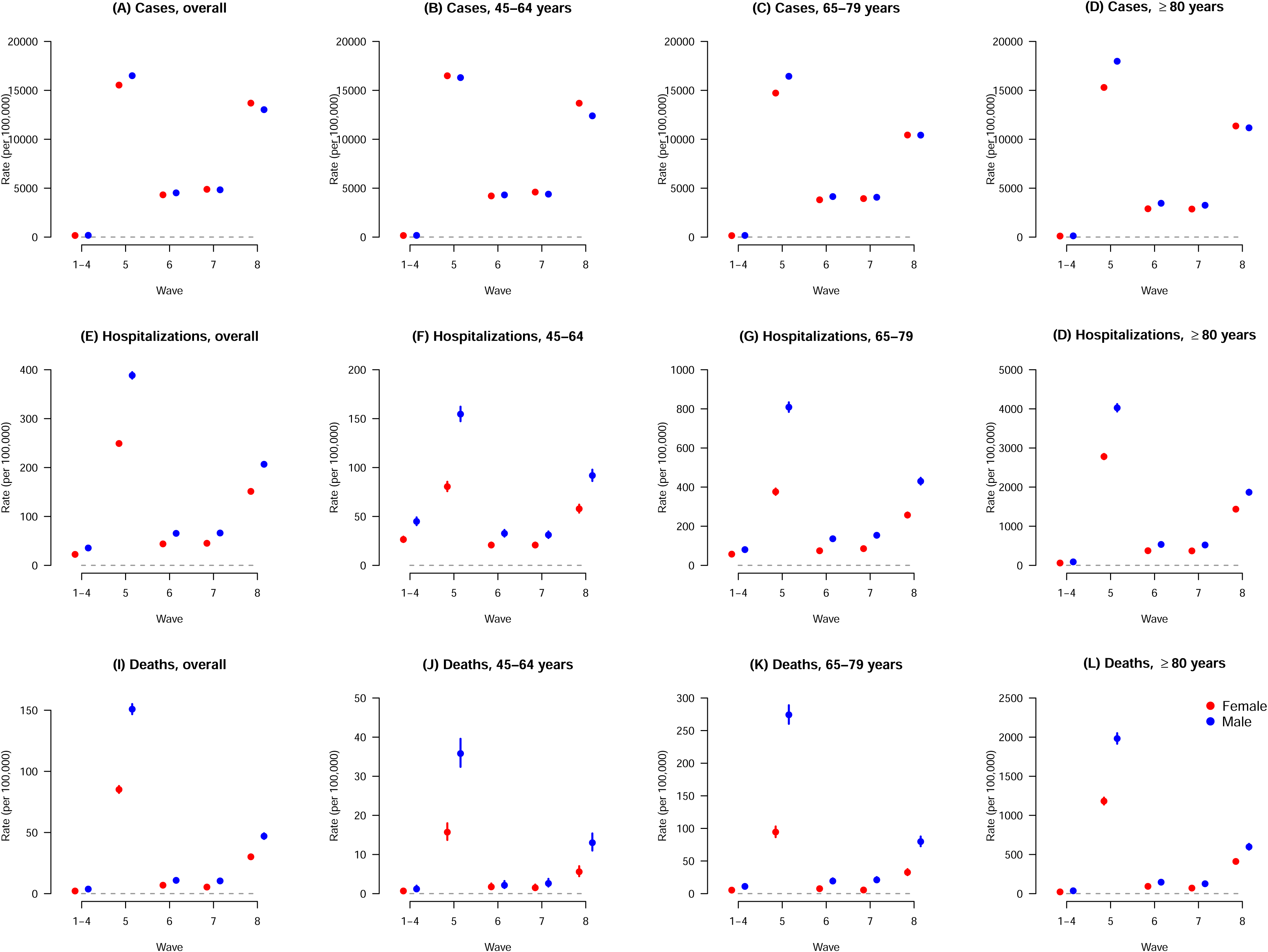
The incidence rates of COVID-19 cases, hospitalizations, and deaths, per 100,000 population (with 95% confidence intervals), by age group and by wave.

Figure 3 presents the estimated sex-, age-, and wave-specific case-hospitalization rate, case-fatality rate (CFR), and hospital-fatality rate. Case hospitalization rates were higher in waves 1-4 than in later waves, and within each wave the rates were similar between males and females. The CFR remained significantly higher for males across all waves, particularly among those aged 80 or above. While the highest number of COVID-19 cases was recorded in wave 5, it did not have the highest CFR. Instead, waves 1-4 show the highest CFR across all ages. The hospital-fatality rate was highest in wave 5, particularly for the population aged 80 or above.

**Figure 3:**
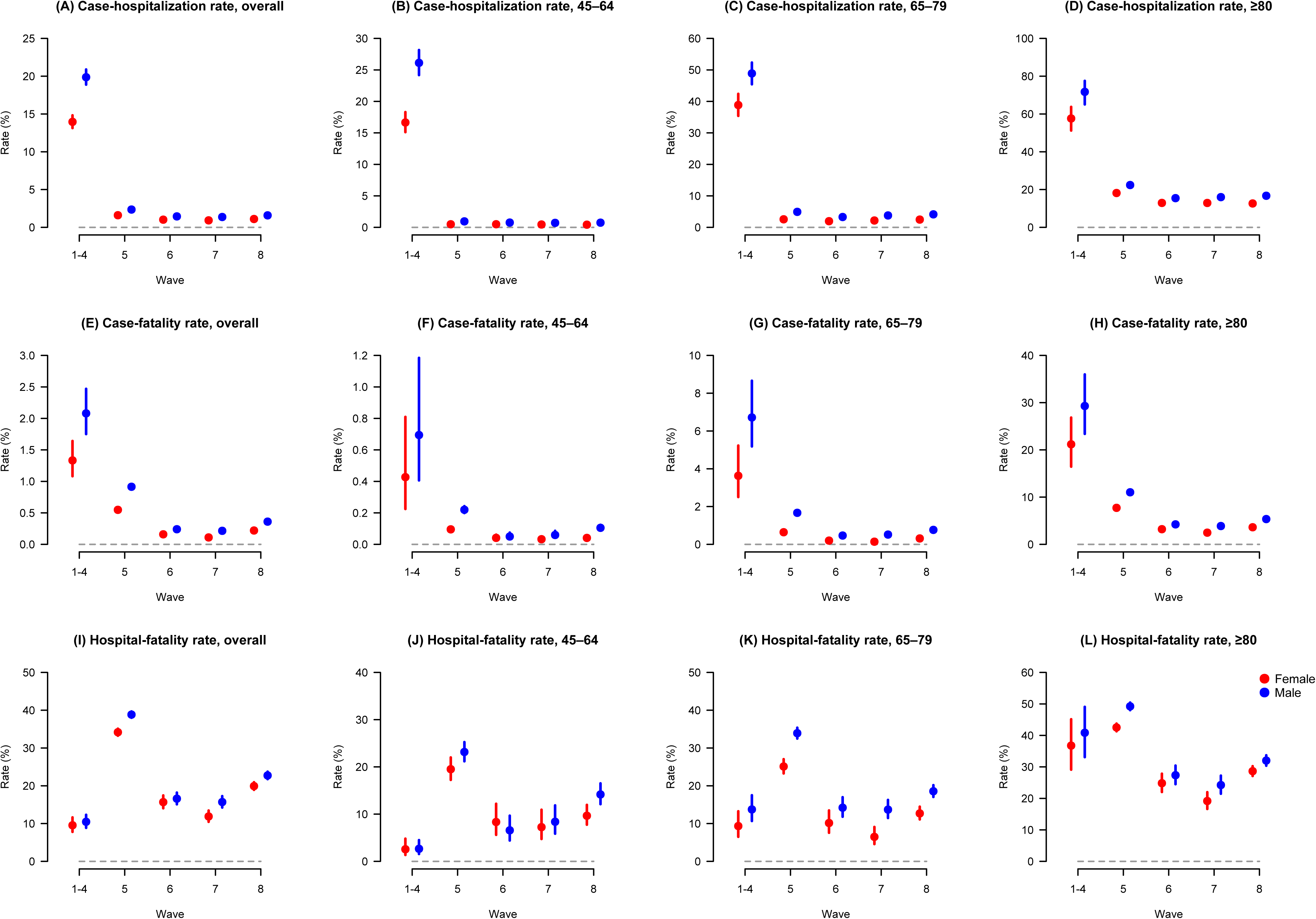
The case-hospitalization risks, case-fatality risks, and hospital-fatality risks for COVID-19 cases per 100,000 population (with 95% confidence intervals), by age group and by wave. Note that some confidence intervals are very narrow and cannot be seen.

Figure 4 presents the estimates of adjusted and unadjusted risks of COVID-19 mortality across age groups. The adjusted model incorporates multiple health conditions, including diabetes, hypertension, obesity, neoplasm, liver disease, kidney disease, upper respiratory issues, chronic obstructive pulmonary disease (COPD), cardiovascular conditions, craniocervical instability, and vaccination status, to assess their effects on mortality.

**Figure 4:**
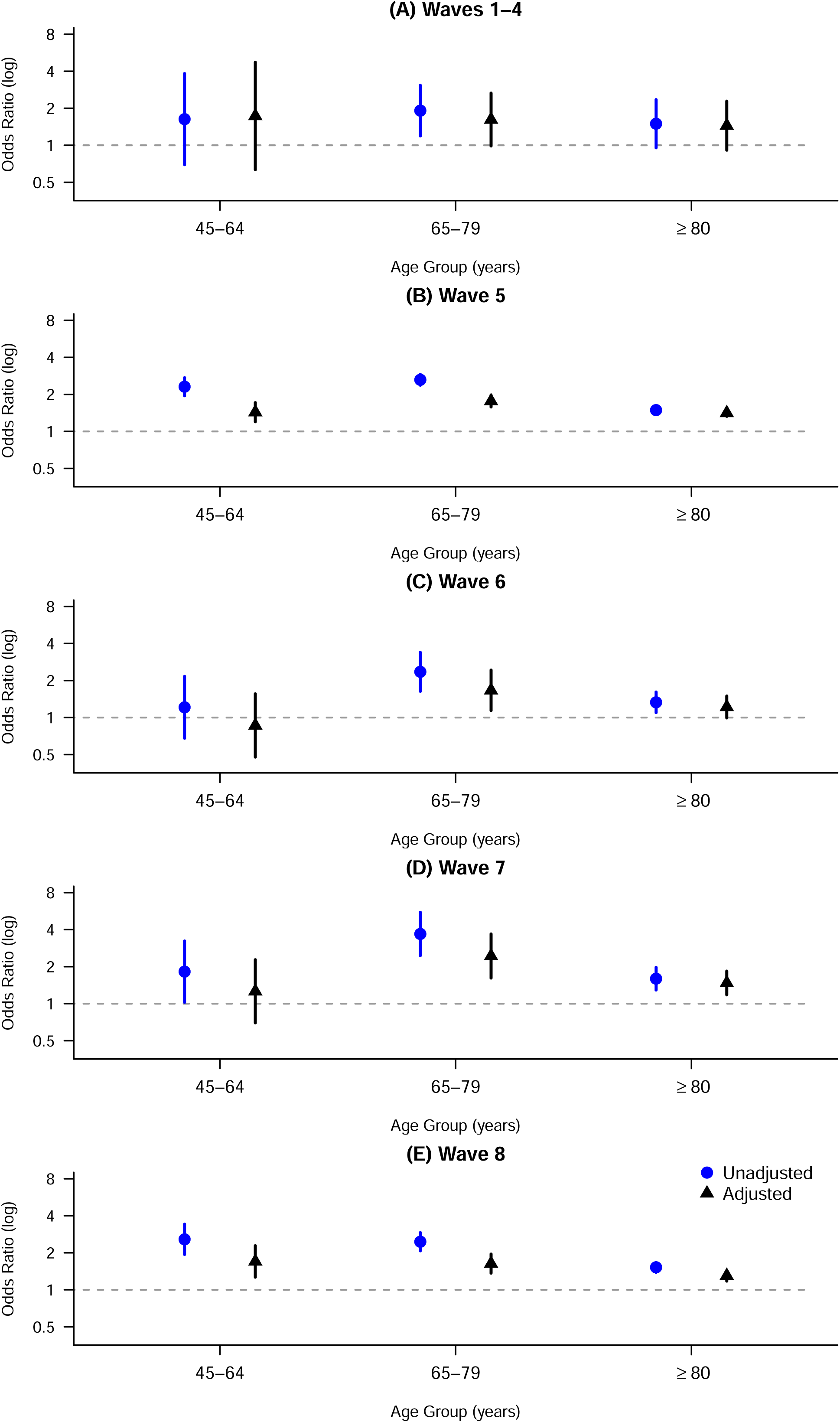
The estimates of the adjusted and unadjusted death odds ratios (with 95% confidence intervals), by epidemic waves and age groups. Note that some confidence intervals are very narrow and cannot be seen.

The unadjusted risks for males are generally higher in the population aged 45 to 64 and 65 to 79 years, throughout all waves. However, the unadjusted and adjusted risks were similar during waves 1-4. The sex difference was particularly pronounced in Wave 5 (adjusted Ors ≈1.4 - 1.8 across age groups) and Wave 7 (adjusted ORs ≈1.5 - 2.4), remaining statistically significant in the majority of age groups even after adjustment. Figure 4 highlights a persistent male excess mortality risk in COVID-19 cases that persisted into later waves. For comparison, we estimated the mortality ratio for respiratory diseases in 2000 through 2019, finding a similar ratio of approximately 1.70 to 2.62 in men (Figure 5).

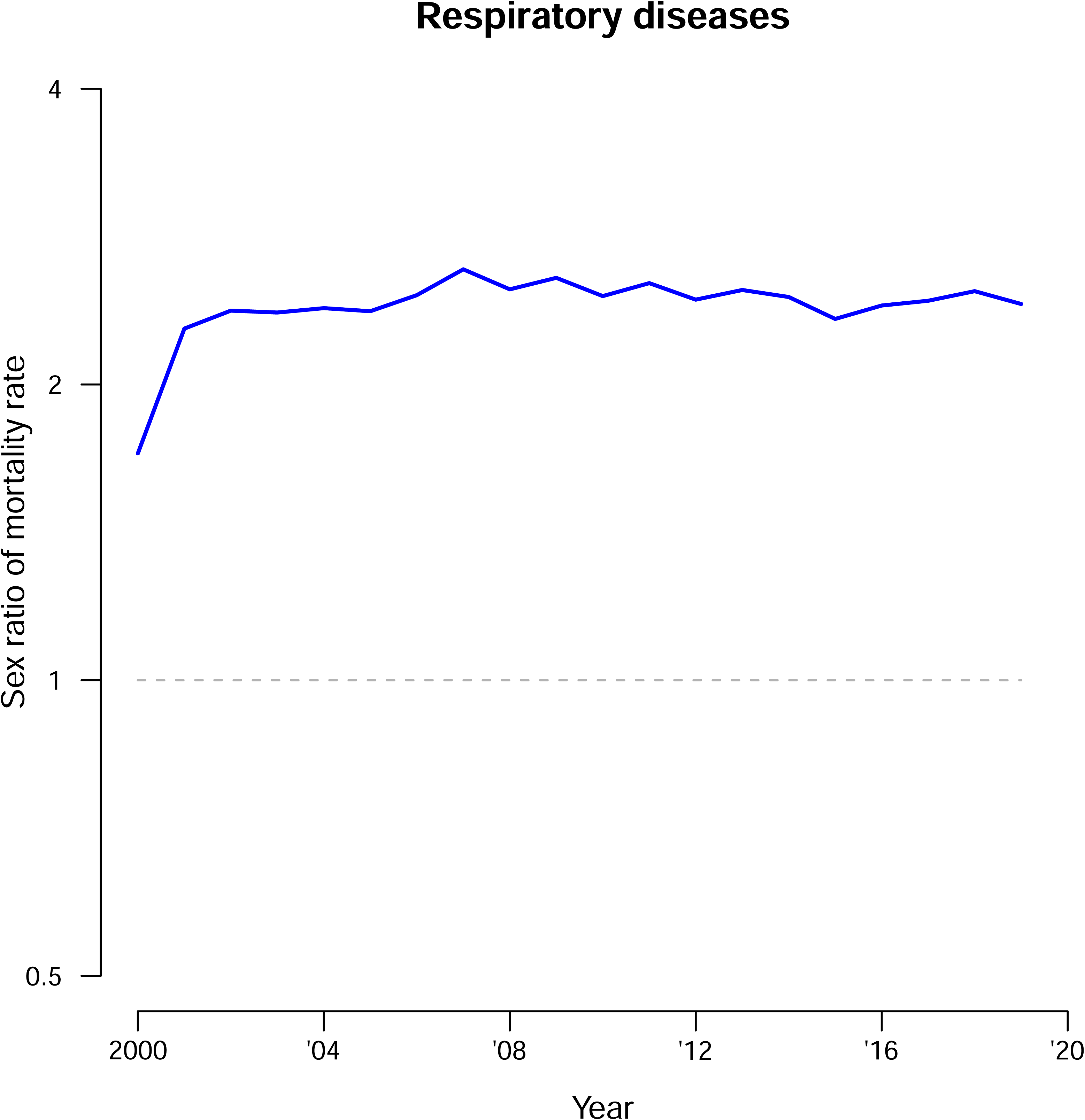

Table 2 presents adjusted odds ratios from logistic regression models predicting COVID-19 mortality. Males had consistently higher odds of death than females across waves (OR 1.44, 95% CI 1.39 - 1.51). Older age was the dominant risk factor, with the highest mortality odds observed among patients aged 80 years and above. The presence of chronic diseases was associated with increased mortality risk overall (OR 1.49, 95% CI 1.39 - 1.59), with variation by wave. Pre-existing kidney disease emerged as one of the strongest risk factors (OR 2.18, 95% CI 2.06 - 2.30), with particularly elevated risk in earlier waves (OR 11.54 in waves 1 - 4). SARS-CoV-2 vaccination had a clear dose-dependent association with reduced mortality, with progressively lower odds ratios for more vaccine doses within each wave.

**Table 2:** The estimated odds ratios and 95% confidence intervals from multivariable logistic regression models predicting COVID-19 mortality among COVID-19 cases.

|  | Odds ratio / Wave |  |  |  |  |  |
| --- | --- | --- | --- | --- | --- | --- |
| Predictor | Overall | 1-4 | 5 | 6 | 7 | 8 |
| Sex - male | 1.44 (1.39 - 1.51) | 1.45 (1.03 - 2.04) | 1.48 (1.40 - 1.56) | 1.23 (1.03 - 1.47) | 1.56 (1.29 - 1.87) | 1.38 (1.27 - 1.51) |
| Chronic diseases | 1.49 (1.39 - 1.59) | 2.06 (1.17 - 3.62) | 1.29 (1.18 - 1.41) | 1.89 (1.40 - 2.56) | 2.16 (1.58 - 2.97) | 1.33 (1.15 - 1.53) |
| Kidney | 2.18 (2.06 - 2.30) | 11.54 (7.09 - 18.76) | 1.87 (1.75 - 2.00) | 2.49 (1.98 - 3.13) | 3.02 (2.39 - 3.82) | 2.34 (2.09 - 2.63) |
| Diabetes | 0.97 (0.92 - 1.03) | 0.89 (0.56 - 1.40) | 1.01 (0.94 - 1.08) | 0.89 (0.70 - 1.13) | 0.64 (0.49 - 0.83) | 0.98 (0.87 - 1.10) |
| Cancer | 1.92 (1.79 - 2.06) | 2.44 (1.27 - 4.69) | 1.62 (1.48 - 1.77) | 2.42 (1.85 - 3.17) | 2.48 (1.88 - 3.26) | 2.19 (1.91 - 2.52) |
| Cardiovascular | 1.61 (1.53 - 1.69) | 2.31 (1.48 - 3.59) | 1.42 (1.33 - 1.51) | 2.08 (1.66 - 2.61) | 1.94 (1.54 - 2.45) | 1.88 (1.68 - 2.10) |
| Hypertension | 0.77 (0.73 - 0.80) | 0.61 (0.39 - 0.95) | 0.82 (0.77 - 0.87) | 0.61 (0.49 - 0.75) | 0.63 (0.50 - 0.79) | 0.73 (0.66 - 0.81) |
| Obesity | 0.71 (0.57 - 0.87) | 0.80 (0.24 - 2.63) | 0.72 (0.55 - 0.94) | 1.01 (0.44 - 2.29) | 0.60 (0.21 - 1.70) | 0.56 (0.35 - 0.89) |
| Upper respiratory | 1.33 (1.06 - 1.65) | 4.60 (1.06 - 20.01) | 1.22 (0.91 - 1.62) | 2.23 (0.95 - 5.24) | 1.35 (0.57 - 3.22) | 1.20 (0.77 - 1.87) |
| Liver | 1.24 (1.04 - 1.46) | 0.72 (0.15 - 3.57) | 1.11 (0.90 - 1.37) | 1.49 (0.78 - 2.86) | 2.32 (1.25 - 4.34) | 1.39 (0.97 - 1.99) |
| COPD | 0.93 (0.86 - 1.01) | 1.02 (0.48 - 2.16) | 0.91 (0.83 - 1.00) | 1.14 (0.83 - 1.57) | 1.04 (0.75 - 1.46) | 0.91 (0.76 - 1.08) |
| Reference Group: No vaccination |  |  |  |  |  |  |
| Vaccine 1st Dose | 0.75 (0.71 - 0.80) | \ | 0.63 (0.59 - 0.68) | 1.12 (0.79 - 1.59) | 1.13 (0.68 - 1.89) | 1.02 (0.79 - 1.32) |
| Vaccine 2nd Dose | 0.46 (0.43 - 0.49) | \ | 0.42 (0.39 - 0.46) | 0.55 (0.44 - 0.68) | 0.79 (0.61 - 1.04) | 0.77 (0.66 - 0.90) |
| Vaccine 3rd Dose | 0.29 (0.27 - 0.30) | \ | 0.22 (0.17 - 0.27) | 0.27 (0.21 - 0.34) | 0.47 (0.37 - 0.59) | 0.47 (0.43 - 0.52) |
| Vaccine 4th Dose | 0.19 (0.16 - 0.21) | \ | \ | 0.11 (0.04 - 0.26) | 0.23 (0.14 - 0.39) | 0.29 (0.25 - 0.34) |
| Reference Group: Age 0-44 |  |  |  |  |  |  |
| Age 45-64 | 9.21 (7.60 - 11.28) | 6.97 (2.03 - 23.98) | 8.57 (6.62 - 11.10) | 10.49 (4.41 - 24.96) | 4.80 (2.44 - 9.44) | 7.62 (5.10 - 11.39) |
| Age 65-79 | 20.31 (16.87 - 24.71) | 48.88 (15.03 - 158.93) | 17.55 (13.67 - 22.53) | 17.88 (7.78 - 41.10) | 7.71 (4.09 - 14.52) | 13.49 (9.18 - 19.84) |
| Age 80+ | 59.34 (49.42 - 72.02) | 272.88 (83.07 - 896.36) | 43.46 (33.95 - 55.63) | 61.92 (27.25 - 140.71) | 25.89 (13.93 - 48.11) | 41.73 (28.55 - 61.00) |

## DISCUSSION

Public health and social measures effectively suppressed the spread of SARS-CoV-2 in Hong Kong for the first two years of the pandemic, but the number of infections and deaths increased dramatically in the fifth wave of the pandemic in early 2022 (11,15,16). While relative infection rates remained comparable between males and females throughout the eight epidemic waves we studied, males experienced consistently higher mortality risks. This aligns with findings of other studies (4,17,18). We found that chronic diseases, including kidney disease, cancer, and cardiovascular disease, were associated with significantly elevated mortality risks, particularly in males. However, after adjustment for the prevalence of chronic conditions, as well as other factors potentially associated with risk of mortality, the adjusted sex ratio remained significantly above 1 (Figure 4).

Our analysis spans the pandemic’s first three years, from low-community-transmission periods, i.e., waves 1-4, to Omicron in waves 5-8 with gradually declining COVID-19 fatality rates. The persistent male excess in mortality, evident in case-fatality rates, hospital-fatality rates, and population-based severity indicators, suggests the sex disparity transcends viral activity levels and likely reflects COVID-19-specific vulnerabilities, including biological factors, that is higher ACE2 receptor expression facilitating viral entry in males (19), and behavioral risks, for example, higher male smoking prevalence of 16.7% vs. 3.0% in females in 2021 (20). Smoking may exacerbate respiratory vulnerability, thus contributing to worse outcomes. Notably, Hong Kong’s coronary heart disease prevalence is higher in males (2.1%) than in females (1.2%) (21), further highlighting sex-specific comorbidity burdens despite equal healthcare access.

Sex differences in COVID-19 infections and mortality have been a common research area for different countries of varied development levels and demographic backgrounds, for effective implementation of interventions, and thus to improve healthcare resource allocation. No significant sex gap was observed in middle-income countries, for example, Poland, Hungary, and Oman (22). However, Hong Kong provides cheap public healthcare with universal coverage, and differential access to care would be unlikely to explain the sex difference in our study.

Apart from exploring the disparity in COVID-19 mortality, sex differences in the mortality that occurred during the COVID-19 pandemic were also explored. Pothisiri et al. suggested that the public health measures in Thailand resulted in lower accidental mortality in men in the early stage of the pandemic because most of the measures, for instance, lockdowns, social distancing, and travel restrictions, might have a greater impact on men than women (23). In Hong Kong, overall mortality rose markedly from 2020 to 2022, with males experiencing a steeper decline in life expectancy in 2022, largely driven by elevated respiratory mortality in older adults, alongside increases in cardiovascular and kidney disease deaths in both sexes (16). These contextual patterns underscore how sex-specific vulnerabilities may interact with pandemic dynamics, complementing the persistent increased risk of mortality in men observed here across waves and age groups (24).

There are several limitations in our study. First, data quality affects the accuracy of our analysis. The inclusion of self-reported RAT-positive cases from the fifth wave onwards could result in biases in case identification if there were differential patterns in self-testing by sex. There could be underreporting of positive cases particularly from the Omicron period onwards when public health measures were being relaxed, and our estimates of incidence rates could be underestimates of the total rates of infections, although we would not expect a major impact on sex differences in rates of COVID-19 hospitalizations and mortality given broad access to laboratory testing in hospitals. We did not model possible sex-specific differences in immune responses or sex-specific behavioral changes in Hong Kong that might also have contributed to sex differences in positive cases and deaths. Finally, our investigation of the sex difference in COVID-19 fatality was limited only to the direct health impact of the pandemic, but did not take the indirect mortality associated with COVID-19 into consideration.

In conclusion, this study explores persistent sex differences in COVID-19 infection and mortality in Hong Kong across three years, finding that males were at higher risk of severe disease even after accounting for key confounders. Shared mechanisms with other respiratory diseases, particularly smoking and comorbidities, likely contribute, highlighting the value of understanding these factors for effective public health responses to current and future infectious disease threats.

## Data Availability

All data produced in the present study are available upon reasonable request to the authors

## ACKNOWLEDGEMENTS

The authors thank Julie Au for administrative support.

## FUNDING

This work was supported by the Research Grants Council of the Hong Kong Special Administrative Region, China (Project No. T11-705/21-N).

## POTENTIAL CONFLICTS OF INTEREST

B.J.C. has consulted for AstraZeneca, Fosun Pharma, GlaxoSmithKline, Haleon, Moderna, Novavax, Pfizer, Roche, Sanofi Pasteur and Seqirus. All other authors report no potential conflicts of interest.

## Notes

### Author Declarations

Our study received ethical approval from the Institutional Review Board of the University of Hong Kong.

