## Appendix for "Sex differences in COVID-19 infection and mortality in Hong Kong"

This appendix provides additional information on the data and statistical methods used within the study.

1. Definition of severe cases based on epidemiological criteria

Clinical records of COVID-19 cases were retrospectively reviewed to define the severity status for patients according to guidelines and local practice in Hong Kong. COVID-19 cases were classified as mild, moderate, severe, critical, and fatal based on ICU admission, condition changes during hospitalization, and oxygen saturation. Drug records during hospitalization were also taken into account to classify severity status, as the choice of treatment was dependent on the cases’ conditions. Table S1 shows the criteria for each classification of cases, and Table S4 shows the proportions of mild, moderate, severe, critical, and fatal cases defined by the proposed criteria in different waves of the pandemic.

Table S1: The criteria for each classification of cases.

| **Case classification** | **Definition** |
| --- | --- |
| **Severity status of cases** | |
| Mild-Moderate | Cases that did not meet the definition of severe, critical, or fatal. |
| Severe | Cases meet either one of the following criteria:  1) On treatment with dexamethasone (+remdesivir), and/or with either baricitinib or IV tocilizumab during hospitalization;  2) Ever being O2 desaturated (O2 desaturation level ≤90%);  3) Ever required an oxygen supplement of 3 litres per minute or more.  Cases that also met the criteria of critical or fatal cases were excluded. |
| Critical | Cases ever admitted to ICU, ever required intubation or extracorporeal membrane oxygenation (ECMO) or in shock. |
| Fatal | For cases in waves 1-4:  A person who died within 28 days of his/her first test positive for COVID-19.  For cases in wave 5a and after:  Recorded in-hospital fatalities by the Hospital Authority. |

Table S2: The proportions of mild-to-moderate, severe, critical, and fatal cases defined by the proposed criteria in each wave of the pandemic, by sex.

|  | Male | | | | | Female | | | | |
| --- | --- | --- | --- | --- | --- | --- | --- | --- | --- | --- |
| Wave | Mild-  moderate | Severe | Critical | Fatal | Total | Mild-  moderate | Severe | Critical | Fatal | Total |
| 1-4 | 4892  (80.1%) | 828  (16.3%) | 258  (4.23%) | 127  (2.08%) | 6105 | 5615 (86%) | 675  (10.3%) | 149  (2.28%) | 87  (1.33%) | 6526 |
| 5 | 544890  (97.6%) | 7220  (1.29%) | 814  (0.15%) | 5102  (0.91%) | 558026 | 616219  (98.4%) | 6039  (0.96%) | 571  (0.09%) | 3432  (0.55%) | 626261 |
| 6 | 150647  (98.6%) | 1639  (1.07%) | 206  (0.13%) | 367  (0.24%) | 152859 | 172259  (99.0%) | 1364  (0.78%) | 126  (0.07%) | 277  (0.16%) | 174026 |
| 7 | 161187  (98.6%) | 1171  (1.05%) | 174  (0.11%) | 351  (0.22%) | 163423 | 194967  (99.1%) | 1475  (0.75%) | 127  (0.065%) | 216  (0.11%) | 196785 |
| 8 | 430025  (98.4%) | 4752  (1.09%) | 604  (0.14%) | 1576  (0.36%) | 436955 | 540576  (98.9%) | 4429  (0.81%) | 404  (0.074%) | 1202  (0.22%) | 546611 |

Table S3: The epidemic wave, period, and predominant variants of each wave of the COVID-19 pandemic in Hong Kong.

| Epidemic wave | Period | Predominant variants |
| --- | --- | --- |
| 1 | 23/1/2020 – 29/2/2020 | Ancestral |
| 2 | 1/3/2020 – 31/5/2020 | Ancestral |
| 3 | 1/6/2020 – 31/10/2020 | Ancestral |
| 4 | 1/11/2020 – 30/12/2021 | Ancestral |
| 5 | 31/12/2021 – 29/4/2022 | Omicron BA.2 |
| 6 | 30/4/2022 – 30/8/2022 | Omicron BA.2 |
| 7 | 31/8/2022 – 26/10/2022 | Omicron XBB |
| 8 | 27/10/2022 – 29/1/2023 | Omicron JN.1 |

2. Definition of COVID-19 cases in Hong Kong

Hong Kong recorded the first COVID-19 cases on 23 January 2020. A wide range of public health and social distancing policies, including contact tracing and compulsory testing for inbound visitors, were implemented to suppress the spread of COVID-19 since 2020. Reverse transcription-quantitative polymerase chain reaction (RT-qPCR) tests were used to detect confirmed cases. Since 26 February 2022, when the number of infections surged, self-report rapid antigen test (RAT) results have been accepted as an alternative confirmation method for COVID-19 cases. Hong Kong has recorded more than 2.9 million locally infected COVID-19 cases, with 1.2 million and 1.6 million cases confirmed with RT-qPCR and RAT, respectively, and 13825 COVID-19 deaths as of 29 May 2023. Table S4 shows the sex-specific distribution of cases confirmed with PCR and RAT tests.

Table S4: The distribution of cases confirmed with PCR and RAT tests in each wave of the pandemic, by sex.

|  | Male | | | | Female | | | |
| --- | --- | --- | --- | --- | --- | --- | --- | --- |
| Wave | Cases (PCR) | Cases (RAT) | Total case | Deaths | Cases (PCR) | Cases (RAT) | Total case | Deaths |
| 1-4 | 6105 | 0 | 6105 | 127 | 6526 | 0 | 6526 | 87 |
| 5 | 366215 | 191811 | 558026 | 5102 | 372738 | 253523 | 626261 | 3432 |
| 6 | 61559 | 91300 | 152859 | 367 | 63831 | 110195 | 174026 | 277 |
| 7 | 50808 | 112615 | 163423 | 351 | 54655 | 142130 | 196785 | 216 |
| 8 | 107027 | 329928 | 436955 | 1576 | 118633 | 427978 | 546611 | 1202 |

Table S5. Hong Kong population by age group and by sex in 2020-2022 (1)

|  | 2020 | | 2021 | | 2022 | |
| --- | --- | --- | --- | --- | --- | --- |
| Age groups | Male | Female | Male | Female | Male | Female |
| 0-11 | 338400 | 318000 | 326300 | 308000 | 315600 | 297100 |
| 12-19 | 229700 | 220300 | 224600 | 213800 | 220700 | 209500 |
| 20-29 | 404500 | 441300 | 379200 | 405400 | 360300 | 378700 |
| 30-39 | 471300 | 679300 | 462500 | 652200 | 452900 | 620300 |
| 40-49 | 479600 | 691200 | 477200 | 692400 | 469400 | 681900 |
| 50-59 | 545800 | 675100 | 529900 | 676200 | 514400 | 668300 |
| 60-69 | 525600 | 545400 | 539800 | 566300 | 557700 | 593900 |
| 70-79 | 261000 | 266300 | 278600 | 289300 | 299900 | 316200 |
| 80+ | 160600 | 227600 | 164200 | 227200 | 163600 | 225700 |
